# Functional changes of visual cortex in pre- and post- operative intermittent exotropia patients: study protocol for a non-randomized case-control clinical trial

**DOI:** 10.1101/2021.11.12.21266258

**Authors:** Yanan Guo, Jing Fu, Jie Hong, Zhaohui Liu, Xueying He

**Author notes:** **Corresponding author:** Jing Fu, No.1, Dong Jiao Min Xiang Street, Dongcheng District, Beijing 100730, China.

## Abstract

**Introduction:** Intermittent exotropia (IXT) is the most common type of divergent squint. IXT is primarily a cortical neurologic dysfunction disorder, occurring as a result of insufficient maintaining of sensory and motor fusion. Recent reports have demonstrated the relationship between IXT and visual cortical impairment. We plan to assess blood oxygen level-dependent (BOLD)- functional magnetic resonance imaging (fMRI) in IXT patients during the pre-and post-operation follow-ups to evaluate the functional changes of the visual cortex.

**Methods and analysis:** A total of 90 Chinese subjects will be recruited, and the age is between 18 and 60 years old. The subjects include the Surgical treatment (ST) group (45 IXT subjects who will perform surgery) and the Healthy control (HC) group (45 age - and sex - and education matched healthy volunteers). The assessments include the following aspects: general ophthalmic, optometry, binocular vision test, Newcastle Control Score (NCS), and fMRI. Each subject completes the rest-state BOLD-fMRI, and the sequences include echo planar imaging (EPI) pulse and 3-dimensional brain volume (3D-BRAVO) to acquire high-resolution images. The follow-up schedule is 6 and 12months after the surgery. The primary outcome will be determined by cortex changes in BOLD-fMRI in ST group before and after the surgery. We also compare the HC group with the pre-operation subjects in the ST group. The secondary outcomes are changes of strabismus examinations, binocular visual function examinations, and NCS.

**Ethics and dissemination:** Ethical approval has been obtained from the Medical Ethics Committee of the Beijing Tongren Hospital. We plan to publish the results of this study in a peer-reviewed journal article.

**Trial registration number:** ChiCTR2100048852

**Strengths and limitations of this study:**

- The present study is advanced which aims to explore the functional changes in the visual cortex in IXT patients before and after surgery through rest-state 3.0T BOLD-fMRI. We also explore the relationships between changes of cortex and ocular examination.
- The follow-up comparison between the visual cortex and ophthalmic changes will enrich our understanding of the impairment and plasticity of visual function in IXT patients.
- Loss to follow-up of participants is possible after the surgery in this study.
- The present study focuses on IXT patients with surgical indications. We did not carry on the study on the different subtypes or the different severity of IXT.

## Introduction

Intermittent exotropia(IXT) is the primary type of exotropia, which is a transitional condition between exophoria and constant exotropia.^1-3^ Few epidemiology articles report the incidence of IXT alone, while instead of reporting the percentage of IXT in exotropia. It is estimated the prevalence of IXT is between 0.8% and 3.5% worldwide among different ages and races, and accounts for over 60% of exotropia.^1-5^ According to the epidemiology reports, the prevalence of IXT is 3.42% to 3.9% in China,^6 7^ and accounts for 50% to 90% exotropia among all age groups.^7-11^

IXT is frequently noted in infancy or early childhood as an outward deviation of one eye. IXT occurred during the disruption of binocular vision, especially when fusional compensatory mechanisms are compromised.^12^ IXT is predominantly a cortical neurologic dysfunction disorder, although with unclear pathology and ecology mechanisms.^13-17^ Along with the disease progression, binocular vision impairment is frequently occurred such as accommodation and vergence deficiency, defective fusion, loss of stereopsis. Gradually IXT breaks down into constant exotropia.^1,6,11-14^ In addition to cosmetic consequences, IXT could have a dramatic impact on education, socialization, and vision-related life quality, if not treated in time.^18 19^ Current treatment strategies include the correction of refractive errors, patching therapy, extraocular muscle surgery, Botulinum toxin injection, and perceptual training, etc.^12^

The fusion function facilitates the brain to translate disparities in information between the images of two eyes into a vergence command to help generate stereopsis, which is a complex cerebral activity, including motor fusion and sensory fusion.^20^ However, fusion is important for orthophoria. Binocular fusion dysfunction possibly disrupts the coordination and balance of the two visual axes, thus leading to misalignment, which is one of the key causes of strabismus.^21^ Surgery is currently the main treatment to align a deviated or strabismic eye for both functional and cosmetic reasons. Weakening the lateral rectus with or without strengthening the medial rectus is commonly used to restore normal eye position. And it was manifested that surgical intervention can restore central fusion and stereoacuity in patients with IXT. Distance stereopsis can recover even if surgery is postponed until adolescence.^22^

Previous studies have found changes and dysfunctions in the cortex in exotropia people, including bilateral medial frontal gyrus, bilateral cerebellum posterior lobe, left angular gyrus and inferior temporal gyrus.^23 24^ Moreover, reports also demonstrated that IXT patients displayed visual cortex dysfunction in bilateral superior parietal lobule and inferior parietal lobule.^25^ However, the exact cerebral region and activation changes during the progression of IXT remain unclear; and the plasticity in the process of treatment needs to be clarified.

Recently, fMRI, especially BOLD-fMRI, has become an important neuroimaging technique used to locate and quantify microscopic functional neurologic changes, with the advantages of well-developed, high-quality, high-resolution, repeatable, and noninvasive.^26^ BOLD-fMRI has been used to evaluate the visual cortex function for decades.^26-28^ Since IXT is related to fusion function and stereopsis impairment, using BOLD-fMRI examination among follow-ups may provide a reasonable method to further reveal the pathological mechanism of IXT.

## Methods

The current prospective, a non-randomized clinical trial is designed to evaluate the functional changes in the visual cortex through rest-state 3.0T BOLD-fMRI during pre-and post-IXT operation, and compare the changes in each time-point during 1-year follow-up. Other changes will also be compared over the study period, including general ophthalmic examinations, optometry examinations, strabismus specialized examinations, binocular visual function examinations, and NCS.

### Study setting and responsibilities

The clinical trial will be conducted in Beijing Tongren Hospital, a large tertiary center with specialist ophthalmology clinics and radiology clinics in Beijing, China. The composition of the steering committee is listed below: Jing Fu is the primary investigator (PI), Zhaohui Liu is the co-PI, Jie Hong is the sub-PI, Yanan Guo, Xueying He are consultants. The steering committee will provide the final approval of the protocol and any changes to the procedure during the clinical trial. Jie Hong will be in charge of supervising the conduction of the study, including staff training and assessment, protocol decisions and amendments, forms development, data management, data analyses, and quality control. Yanan Guo and Xueying He are responsible for data collecting and recording.

### Study design and recruitment

This study has been designed as a non-randomized case-control clinical trial. The recruitment will officially begin on September 1st, 2021. A total of 90 will be recruited, and each of the participants in the ST group will be followed for 1 year.

Potential participants will be recruited for the clinical trials in Tongren Hospital through two primary processes: (1) an ophthalmologist referral during daily routine outpatient clinical work; and (2) an optometrist referral from optometry clinics during myopia treatment. All of the potentially eligible participants will be contacted by a study staff who will explain the process of the study in detail to ensure that patients understand the entire clinical trial. Prior to signing informed consent, detailed information about the study will be provided to participants, including the purpose of the research, examinations, follow-up duration, and possible risks. If they are interested, the subjects will be seen in the clinic to sign the informed consent form. All the identifiable information is confidential.

Intended subjects will be invited for the eligibility and baseline assessments by the research staff. Staff screens patients who meet the inclusion criteria in the strabismus ophthalmology clinic. As soon as finishing the exams and meeting the inclusion criteria, eligible subjects are enrolled. The flowchart of the clinical trial is shown in Figure 1.

**Figure 1.**
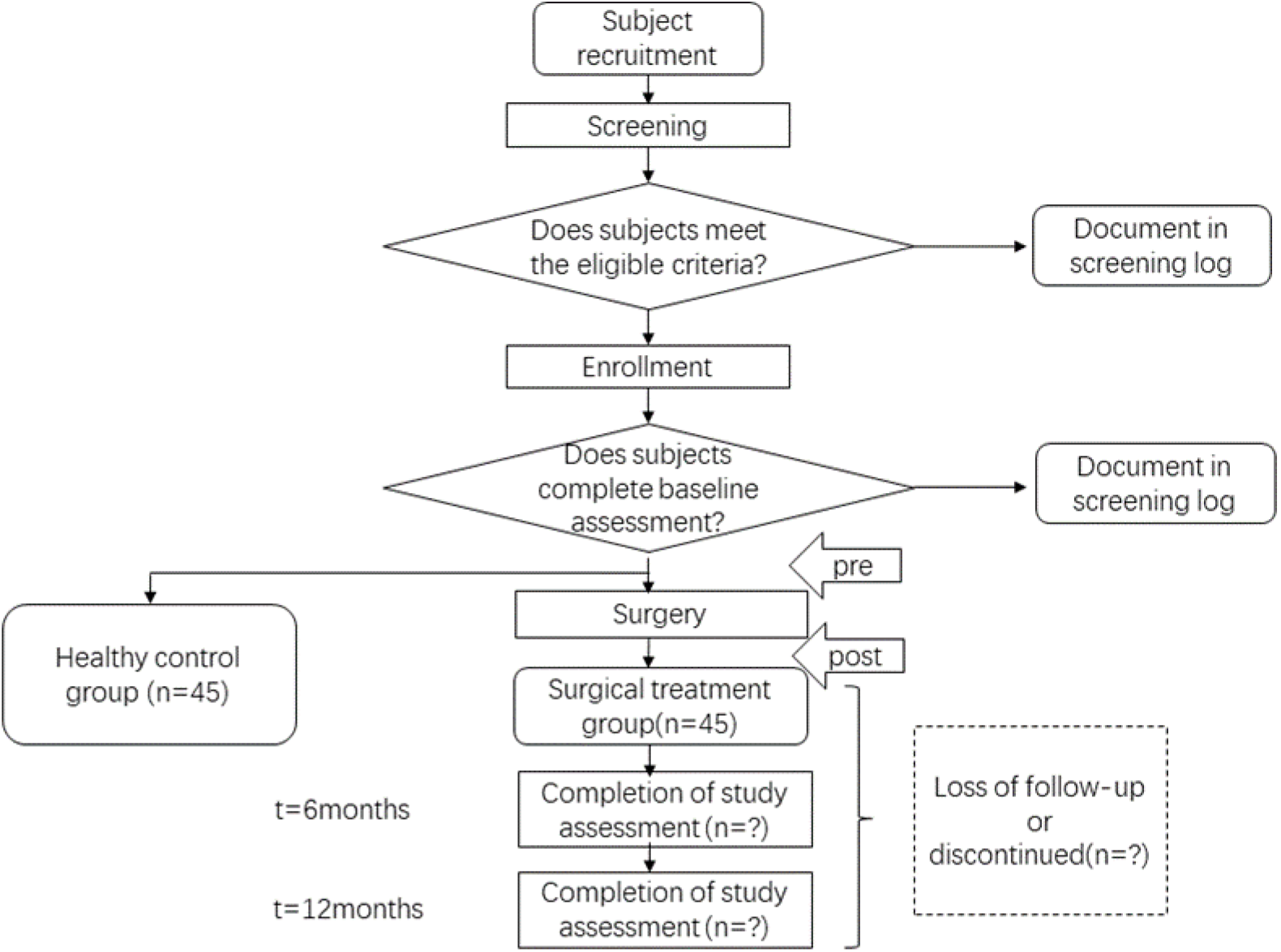
Schematic of the trial design.

Once the subjects are enrolled, retention efforts will be addressed to participants. The Staff of the study will (1) provide periodic contacts about the IXT situation of the subjects; (2) provide feedback regarding the eye conditions, and (3) make an appointment for the next review and follow-up visits.

### Participants and sample size

Potential participants will contact a study staff and will be provided with an information checklist. Interested participants will complete the clinic screening and, if appropriate, will be invited for an eligibility and baseline assessment by the study staff. The patient provides written informed consent for participation in the study prior to any trial-specific procedures.

Estimation of sample size is based on the two following methods: statistical analysis and sample size from previous articles.

Statistical analysis: The two groups of independent sample rate comparison were used for the sample size calculation. According to a previously published article, the improvement rate in distance stereopsis of IXT patients after surgery was 28%.^29^ And the rate of HCs was supposed to be 0. Other parameters that were used include α=0.05, 1-β=0.8, two-sided and 1:1 allocation to the two study groups. For the above parameters, 28% improvement in the ST group vs 0 % in the HC group to be detected at a significance level of p< 0.05, and the estimated sample size for each group is 21 subjects. Based on our previous experience with clinical trials for treating IXT, we estimated the rate of subject loss to follow-up over 1 year to be approximately 50%. Considering these factors together, the estimated sample size for each group is 42.

Sample size from previous articles: Key information about the sample size calculation is missing in published articles related to strabismus, IXT, and fMRI, and a reasonable number of subjects per group ranges from 5 to 32 without follow-up.^21 23 30-35^

Based on previous articles and the statistical calculation, a sample of 45 eligible subjects will be required in the ST group of the trial. To compare the changes in the cerebral area, normal controls are also recruited. A total of 45 healthy age-matched HCs who meet the entry criteria will be enrolled.

### Eligibility criteria

Inclusion criteria:

1. Age between 18 and 40 years;^21 23 31 33-35^
2. Evidence of IXT on the basis of history and clinical examination;
3. Have surgical intention and meet the surgical indications including loss of distant stereopsis using synoptophore and/or deflection time accounts for more than half of waking time; angle of deviation< -15^△^;
4. No ongoing or planned amblyopia treatment;
5. Best-corrected visual acuity better than or equal to 20/20 in both eyes, no anisometropia (< 1.50 D and < 1.00 D in the difference in spherical and cylindrical error, respectively)^36^;
6. Right handedness;
7. Be able to understand and cooperate with examinations, sign informed consent voluntarily.

HCs are recruited based on the NO.1, 4, 5, 6, and 7 in the inclusion criteria.

Exclusion criteria (Subjects will be excluded if they meet any of the following criteria):

1. Structural ocular pathology;
2. Previous eye surgery history;
3. Accompanied by vertical strabismus;
4. History of diseases of the central nervous system and the whole body;
5. History of psychiatric diseases including depressive disorder and delusional disorder;
6. Contraindication for MRI examination (treatable or accidental magnetizable metal in cardiac pacemaker or prosthesis or previous head or spinal trauma requiring neurosurgery, etc.);
7. Claustrophobia. The study center is a tertiary A hospital. A chief physician(JF) performs the examination of strabismus and the surgery, and a deputy chief radiologist (ZHL) performs fMRI examination. After the surgery, patients whose deviation is less than 10 prism diopters are included.^37^

### Examinations and Study outcomes

IXT is presented as a divergent misalignment of the visual axes. The deviation often becomes manifest with fatigue, visual inattention, or illness when fusional compensatory mechanisms are compromised.^12^ Previous studies have found changes in the visual cortex in IXT patients. In addition, the deviation degree, NCS, and several influencing factors are also related to the evaluation of IXT. Taken together, in the present project, the primary, secondary and exploratory outcomes will be evaluated during the 12 months follow-up period according to the schedule as follows (Table 1 and Figure 1).

**Table1.** Schedule of assessments and outcomes items

| Procedures/Measurements |  | Enrollment*<br>(-2 to 0 week) | Baseline | 6 months<br>(±14 days) | 12 months<br>(±21 days) |
| --- | --- | --- | --- | --- | --- |
| Consent form signed |  |  | × | × | × |
| Basic information | Demography |  |  |  |  |
|  | History |  |  | × | × |
| Refraction | Noncycloplegic<br>autorefraction |  |  | × | × |
|  | Cycloplegic subjective<br>refraction |  |  | × | × |
|  | Cycloplegic<br>autorefraction |  |  | × | × |
| Visual acuity | Best corrected visual<br>acuity |  |  | × | × |
| Strabismus<br>examinations | Ocular dominance |  |  |  |  |
|  | Cover test (distance,<br>near) |  |  |  |  |
|  | Prism cover test<br>(distance, near) |  |  |  |  |
|  | Ocular movements |  |  |  |  |
|  | Distance stereopsis<br>(synoptophore) |  |  |  |  |
|  | AC/A ratio |  |  |  |  |
|  | Distance stereopsis<br>(random-dot<br>stereogram) |  |  |  |  |
|  | Visual perception<br>examination |  |  |  |  |
|  | NCS evaluation |  |  |  |  |
| Eye examinations | Slit-lamp exam |  |  | × | × |
|  | Retinal photography |  |  | × | × |
| fMRI | Rest-state | × |  | × |  |

\*Enrollment time-point could be the same day of baseline measurement.

### Primary outcome

rest-state BOLD-fMRI detection is performed immediately after recruitment as baseline data and will be evaluated in the 1-year follow-up. For the primary outcome analyses, the changes between pre-operation and post-operation will be determined by the values between baseline and the last follow-up visit (1 year after the surgery). Other measurements obtained at each follow-up visit are considered secondary outcomes measures.

### Procedures for fMRI detection

After recruitment, an appointment for fMRI examination will be registered by research staff. Rest-state 3.0T magnetic resonance scanner (General Electric Medical Systems, Milwaukee, WI, USA) is used to scan the visual cortex. Matched eight-channel phased array coil with earplugs and foam padding will be used to lessen scanner noise and head-motion. Rest-state fMRI is running for 40 minutes. Subjects are asked to close their eyes, stay awake.

Rest-state fMRI: An EPI pulse sequence is used, and imaging protocols are as follows: a repetition time (TR)/ echo time (TE) = 2000/35 ms, flip angle = 90°, field of view (FOV) = 240mm × 240mm, matrix = 64 × 64. 28 axial slices are obtained with 4mm thickness and a 1mm gap. In each fMRI session, the pulse duration is 400 seconds, and 3D-BRAVO sequences are used to acquire high-resolution structural images (TR = 8.8ms, TE = 3.5ms, TI = 450ms, matrix = 256 × 256, FOV = 240mm × 240mm, slice thickness = 1.0mm without gap, flip angle=13°). During the scan, participants are required to stay motionless, stay awake and stay focused until the scan is over.

### Secondary outcome

The following binocular vision test are undertaken at each visit (Table 1), to assess the basic ocular conditions, the severity of strabismus, and the extent of damage to stereopsis.

Ocular dominance: subjects are asked to hold a card with a central hole and fixate on a distant object while holding their head stationary. The examiner covers the subjects’ eyes one after the other and repeats three times to determine the ocular dominance eye. If the results are inconsistent, the dominant eye is recorded as indefinite.

Cover test: cover test is used to assess the presence and magnitude of heterophoria and strabismus by asking the subject to fixate on targets at 33 cm and 6 m, with and without spectacles, if worn, conducted by an experienced ophthalmologist. The Hirschberg test is performed to screen the presence or absence of strabismus. If strabismus is present, the cover-uncover test is used to differentiate phorias and tropias, to determine if the tropia is intermittent or constant, and to differentiate unilaterally (right or left) and alternating tropia. If no strabismus is detected, the alternating cover test is performed to detect heterophoria.

Prism cover test: subjects with strabismus are further measured in prism diopters in the alternating cover test using loose prisms (GZS-01TYPE, Tianyuehengtong Medical Co. Ltd, Tianjin, China). The prism is placed with the base along the reversed direction of deviation and is adjusted until no movement could be detected.

Ocular movements: to record the ocular motility, nine directions of gaze including primary (straight ahead), secondary (right, up, left and down), and tertiary (upper right, lower right, upper left, and lower left) are examined and recorded by asking subjects fixating on a moving penlight without moving their heads.

Synoptophore (2001, Clement Clarke, UK) is used to observe the far distance stereopsis, accommodative convergence/accommodation (AC/A) ratio. Random-dot stereogram designed by Yan Shaoming and visual perception examination to check the function of near stereopsis is performed. Visual perception examination includes static stereopsis, perceptual eye position, and dynamic stereopsis. It performs on a Windows XP computer mainframe and LG2342p polarized three-dimensional displayer. The resolution of the displayer is 1920 ×1080 and the refresh frequency is 120Hz. The visual perception inspection and evaluation system are developed by the national research center of medical and health appliance engineering technology. During the examination, subjects take the sitting position, with their eyes 0.8m away from the center point of the monitor and at the same height as the monitor. They wear polarized glasses for binocular separation and respond to the examination by mouse or keyboard.

NCS is used to assess how well the squint is controlled by IXT subjects.^38^ The NCS combines an estimate of the observed frequency of the IXT (home control) with an assessment of the subject’s ability to realign the eye following a cover test to induce misalignment (Table 2).

**Table2.**
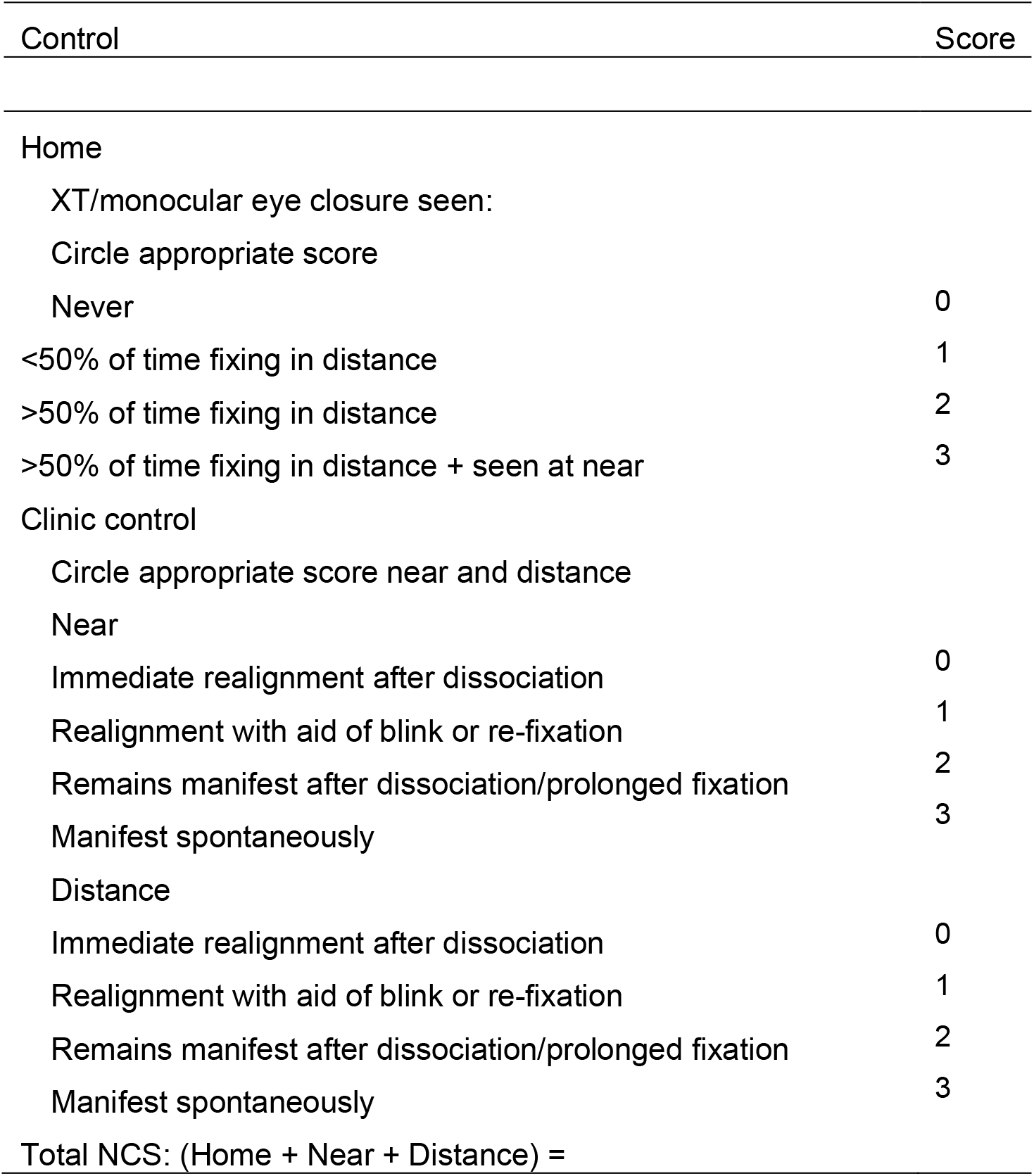
Newcastle Control Score for intermittent exotropia (revised)

### Exploratory outcomes

1. General ophthalmic examinations: slit lamp biomicroscope (SL-3G, Topcon, Tokyo, Japan) and retinal photography (TRC-50DX/TRC-50DX, Topcon, Tokyo, Japan) would be performed to identify any abnormalities of the eye, including anterior segment, refractive media, and fundus examination.
2. BCVA: corrected vision at a distance of 6m is examined using LogMAR visual acuity chart.
3. Optometry examination: 2 drops of 1% tropicamide are given 5 minutes separately to dilate the pupils. Cycloplegic refraction outcome measures will be obtained 30-45 minutes after the first drop is instilled, which ensures the maximal cycloplegic effect. Objective refraction is measured before and after cycloplegia using an autorefractor (KR-800, Topcon, Tokyo, Japan) followed by subjective refraction by trained optometrists.

### Follow-up examinations and measurement schedule

The exanimation measures will be assessed at enrollment/baseline, 6 months, 12 months. The differences in all of the mean values at each follow-up visit from baseline will be analyzed (Figure 1).

### Interventions and Allocation

#### Surgical treatment (ST) group

IXT patients who have a definite diagnosis and conform to surgical indications and agree to surgical treatment are included. All of the patients will be re-examined according to the schedule (Table 1 and Figure 1). Before each follow-up visit, the coordinator will make an appointment in advance.

#### Healthy control (HC) group

We recruit age - and sex- and education-matched volunteers to serve as the HC group. The HC group is set up to find out brain differences between healthy people and IXT patients.

Ophthalmologists will provide advise on the operation choices according to the subjects’ condition and treatment guidelines. The patients will freely choose the treatment methods, including whether to have the operation or not. Patients who have both surgical indications and surgical intentions are enrolled in the present study. So randomization is not applicable for this trial. Whether the patient chooses to participate in this study will not influence his/her next step in treatment.

### Patient and public involvement

Patients and/or the public were not involved in the design, or conduct, or reporting, or dissemination plans of our research.

### Statistical methods

#### Data management and data analyses

For the quantitative data, tests for normality and homogeneity of variance are performed first. Quantitative data conforming to normal distribution are described as mean ±standard deviation. The difference between the ST group and the HC group is compared by a two-sample *t*-test. The difference between the baseline and 12 months after the surgery in the ST group is compared by paired *t* test. The quantitative data of skewness distribution are described by Median (Q25,Q75). Mann-Whitney U test is used to compare the difference between the ST group and the HC group, and Wilcoxon signed-rank test is used to compare the difference between the baseline and 12 months after surgery in the ST group. Qualitative data are described as proportion, and the difference between groups is compared by chi-square test or Fisher’s exact test. P < 0.05 is considered statistically significant.

#### General information

Age from the two groups will be presented as the mean ± standard deviation. Gender and subject number in each group will be presented as proportion.

#### Results of fMRI

The preprocessing is performed using Data Processing Assistant for Rest-State fMRI (DPARSF 2.1; State Key Laboratory of Cognitive Neuroscience and Learning, Beijing Normal University, Beijing, China; available in the public domain at http://restfmri.net/forum/DPARSF). It is based on Statistical Parametric Mapping (SPM12) (http://www.fil.ion.ucl.ac.uk/spm/) running by MATLAB R2013b (The MathWorks, Natick, USA). The first 10 volumes will be removed in order that participants to adapt to the scanning noise and the signal equilibrium after converting DICOM files to NIFTI images. Next, we will perform slice timing, head motion correction, spatial normalization to the Montreal Neurological Institute template (resampling voxel size = 3mm × 3mm × 3 mm), linear trend removal, temporally bandpass filtering (0.01–0.08Hz), and spatially smoothed with a Gaussian kernel of 4mm full-width at half-maximum. If the subject’s head is shifted by more than 1.5mm or rotated by more than 1.5°during the treatment, the subject’s data will be discarded. The amplitude of low-frequency fluctuation (ALFF) and fractional amplitude of low-frequency fluctuation (fALFF) values are calculated using DPARSF. The fALFF is calculated for slow 4(0.027–0.073 Hz) and slow 5(0.01–0.027 Hz) bands.

#### Results of ocular examination

deviation degree, AC/A ratio, near stereopsis, NCS, static stereopsis, and perceptual eye position in visual perception examination are described as mean ± standard deviation. Far stereopsis, dynamic stereopsis in visual perception examination are described as proportion. Pearson correlation analysis is performed to explore the relationships between mean ALFF/fALFF values of discrepant brain regions and ocular examination in the patient group.

#### Oversight and monitoring

During the study, data will be entered into a secure database with range checks for each data field. Anonymized paper copies of forms will be stored in secure locked file cabinets. The final dataset will be used after approval by the Steering Committee. Medical records will be kept intact in the hospital. The staff will record the results of the examination on the subject’s medical records. The investigator and the ethics committee will be allowed to review the subject’s medical records.

Participants can withdraw from the trial without giving a reason. In addition, investigators may also withdraw them from the study to protect participants’ safety. Any public report of the results of this study will not disclose the subject’s personal identity. We will make efforts to protect the privacy of personal medical data to the extent permitted by law. In accordance with medical research ethics, in addition to personal privacy information, trial data will be available for public inquiry and sharing, which will be limited to web-based electronic databases to ensure that no personal privacy information is disclosed.

We will establish an internal data monitoring committee (DMC) which will consist of ophthalmologists who do not participate in the running trial, statistical experts, and ethics committee members. Dr. Yan Liu will be the chair of DMC. DMC will perform data monitoring quarterly, including monitoring safety information, data completeness, and adverse events, etc. The integrity of the trial for each participant will be checked to ensure the appropriate allocation, completeness of outcomes, accuracy, and timeliness of data collection, etc. We will select an independent monitor to check all signed Consent Forms to ensure the validity. All the examinations are general items in the daily work and are non-invasive. The trial has low risk and no Data Safety Monitoring Committee is established for the present trial.

The present study was approved by the Medical Ethics Committee of Beijing Tongren Hospital and conformed to all the principles required by the Declaration of Helsinki.

#### Potential risks and adverse events

This is an observational low-risk trial and risks could come from two aspects. Firstly, the duration of fMRI examination is long, and the average head scan takes about 40 minutes. There is obvious noise in the scanning chamber. Secondly, MRI is sensitive to the movement of subjects and easy to produce artifacts, so the subject can’t move during the scan.

If there is any discomfort or any unexpected circumstances occured during the study period, whether related to the study or not, we will terminate the examination as soon as possible. The participants can withdraw from the study without giving a reason. Doctors will do their best to prevent and treat possible injuries from this study. If a participant is injured due to the participation of this study, we will provide the necessary medical treatment. According to relevant laws and regulations in China, the research group bears the corresponding medical expenses and provides the corresponding financial compensation.

## Discussion

### Binocular vision and IXT occurrence and development

Binocular fusion includes simultaneous perception, flat fusion, and stereopsis according to Worth’s classification.^39^ Approximately 70% of the cells in the striate cortex are binocular cells, any impairment of fusion and higher spatial perception is closely related to the course of IXT. Fusion function, including sensory fusion and motor fusion, involved V1, V2, V3, MT region, and higher-level ventral and dorsal visual pathways coordination.^40 41^ Sensory fusion is a cortical process that is defined as the unification of visual excitations from corresponding retinal images into a single binocular stereoscopic visual percept. Motor fusion refers to the ability that allows fine-tuning of eye position to align the eyes and maintain eye alignment, which in such a manner that sensory fusion can be maintained.^42 43^ Any abnormal visual experience, such as anisometropia, ametropia, form-deprivation, could destroy binocular fusion by affecting binocular neurons.^14^ The abnormal function of binocular fusion will destroy the coordination and balance of the two visual axes. Once the ocular axes separate, visual feedback has no impact on the subsequent outward movement of the deviating eye to its final position, thus leading to eye malposition. After fusion loss, the deviating eye moves outward in a stereotypic, reflexive fashion.^44^ Xue and his colleagues reported that preoperative distance stereopsis and fusion of most IXT patients were damaged. Six weeks postoperatively, all patients exhibited peripheral fusion and 98% demonstrated central fusion compared to 94% and 40% preoperatively. And the percentage of distance stereoscopic improvement was 77% compared to 13% preoperatively.^22^ Strabismus surgery in adults has been often regarded as merely cosmetic. But the ideal goal of the surgery is to restore binocular function and stereopsis as well as motor alignment,^37^ which could improve health-related quality of life in adults. Recently, several studies have shown improved sensory status and quality of life in adult patients after successful strabismus surgery. Some adults may also regain fusion and stereopsis and increase the field of binocular vision, suggesting that there may be residual neural plasticity in adulthood. ^45-48^

Previous studies have demonstrated the exact visual cortex area of the control center of fusion and showed the changes of cerebral in exotropia patients. Yang et al. reported the bilateral frontal gyrus and left lingual visual cortex regulate normal fusion function in human eyes.^21^ Zhu reported the concomitant exotropia patients exhibited significantly less functional connectivity (FC) between the left Brodmann Area (BA) and left lingual gyrus/cerebellum posterior lobe, right middle occipital gyrus (MOG), left precentral gyrus/postcentral gyrus and right inferior parietal lobule/postcentral gyrus, and significantly less FC between right BA17 and right MOG, which may underlie the pathologic mechanism of impaired fusion and stereopsis in concomitant exotropia patients.^34^ Shi et al. reported in patients with constant exotropia, the right V2 showed increased regional homogeneity (ReHo) values, whereas the left BA47 demonstrated decreased spontaneous ReHo values compared with HCs.^35^ Li et al. demonstrated the fractional anisotropy values in the right inferior frontal-occipital fasciculus (FOF) and right inferior longitudinal fasciculus were significantly higher and the radial diffusivity values in the bilateral FOF, forceps minor, left anterior corona radiata, and left anterior thalamic radiation were significantly lower in the comitant exotropia group than in the HCs.^31^ With voxel-based morphometry, smaller volumes of gray matter were found in the occipital eye field and parietal eye field, and greater volumes in the frontal eye field, supplementary eye field, and prefrontal cortex in concomitant exotropia.^49^ Many areas in the cortex and/or subcortical levels of IXT patients also have abnormal functions. Li et al. reported the increased activation intensity was observed in bilateral MOG, right inferior temporal lobule, right precuneus and fusiform gyrus, and lingual gyrus in the right occipital lobe in subjects with IXT compared with normal subjects.^25^

Overall, assessing the visual cortex of patients with IXT is able to evaluate changes related to stereopsis and other functions. However, there are few studies that focus on the plasticity of IXT patients between cerebral fusion dysfunction and ophthalmic outcomes in follow-ups. In our study, we perform ophthalmic examinations such as binocular visual function, NCS as well as BOLD-fMRI during the follow-up pre-and post-operation, which can help us to analyze the changes of fusion function in the eyes and visual cortex comprehensively and provide evidence for the correlation of IXT and cerebral unbalanced activation. Whether the fMRI changes are related to the binocular visual function changes or not remains unknown before the trial is finished. Maybe cortex changes fail to improve binocular visual function. It indicates that the changes of the cortex are not synchronized with the changes of binocular visual function, or that the slight changes of the cortex are not enough to cause the changes of binocular visual function, or that the existing binocular visual test is not sensitive enough. The results might provide clues to find out potential explanations to the role the central nervous system plays in the pathogenesis and ecology of IXT.

### Rationale of the study design

In recent years, fMRI has become a useful tool for exploring the central mechanism of fusion function as a non-invasive assessment method, which probably can be combined with clinical examination to improve the diagnosis and treatment of IXT. The main advantages of fMRI are high spatial resolution and the ability to reveal detailed microscopic structural changes.^26^ BOLD-fMRI uses the principle of blood oxygenation level dependence, that is, inconsistencies in the local hemodynamics of neurons following excitation, in order to reveal spontaneous neuronal activity by quantifying alterations in blood oxygen level signals ^50^. Besides, 3.0T fMRI has better signal-noise and contrast-noise compared to clinically used conventional 1.5T fMRI. Thus, the present study will employ the 3.0T BOLD-fMRI to localize and quantify brain perceptual activities.

Furthermore, rest-state fMRI is an instrumental research method in vision-induced fusion function evaluation. Previously, rest-state fMRI is widely used to evaluate the changes in the visual cortex. In our preliminary pre-experimental study, we measured the amplitude of low-frequency fluctuation (ALFF) values and fractional amplitude of low-frequency fluctuation (fALFF) values using rest-state fMRI. Compared with HCs, ALFF/fALFF values significantly increased in the bilateral inferior parietal lobe, MOG, and inferior frontal gyrus, and decreased in bilateral postcentral gyrus, precuneus gyrus, and right precentral gyrus of IXT patients. Functional examinations of the visual cortex combined with IXT perioperative evaluation have provided a beneficial exploration, such as further research about the correlation between visual cortex impairment, plasticity, pathogenesis, treatment, and prognosis in IXT patients.

### Potential findings and significance

Nonhuman primates prevented the two eyes from drifting apart by regulating the cortical binocular mechanisms, which maintain fusion. When one eye is occluded, the cortical drive is interrupted, and the covered eye drifts outward.^51^ As is shown in our overall study schedule, we evaluate the changes of the visual cortex before and after the IXT operation and aim to find the connection between binocular visual function and visual cortex function in IXT patients. Meanwhile, the findings could provide a non-invasive method for the assessment of visual function via rest-state BOLD-fMRI, which may be potentially conducted as an evaluation method in patients with strabismus and amblyopia.

### Trial status

The ethical approval has been proven by Tongren Hospital, Capital Medical University (TRECKY2019-147; version number: v1.0; date: 2019-12-11). Participant recruitment will start on 2021-09-01, and the approximate date when recruitment will be completed by 2023-8-31.

## Supporting information

Checklist

## Data Availability

After the clinical trial is finished, the original data will be uploaded to the ResMan Primitive Data Sharing Platform (IPD Sharing Platform) of the China Clinical Trials Registry at http://wwww.medresman.org:22280/login.aspx.

## List of abbreviations

AC/A: accommodative convergence/accommodation
ALFF: amplitude of low-frequency fluctuation
BCVA: best corrected visual acuity
BOLD: blood oxygen level-dependent
BA: Brodmann area
DMC: data monitoring committee
DPARSF: Data Processing Assistant for Rest-State fMRI
EPI: echo planar imaging
fALFF: fractional amplitude of low-frequency fluctuation
FC: functional connectivity
fMRI: functional magnetic resonance imaging
FOF: fronto-occipital fasciculus
FOV: field of view
HC: healthy control
IXT: intermittent exotropia
MOG: middle occipital gyrus
NCS: Newcastle Control Score
PI: primary investigator
ReHo: regional homogeneity
SPM: statistical Parametric Mapping
ST: surgical treatment
TE: echo time
TR: repetition time
3D-BRAVO: 3-dimensional brain volume

## Declarations

### Ethical approval and consent to participate

Ethical approval has been obtained from the Medical Ethics Committee of the Beijing Tongren Hospital, and the study protocol follows the principles described in the Declaration of Helsinki for research involving human subjects. Informed consent will be obtained from all participants prior to their inclusion in the study. In each procedure during the clinical trial, all of the participants should be involved. Patient recruitment will start on 2021-09-01.

### Patient consent for publication

Obtained.

### Provenance and peer review

Not commissioned; externally peer reviewed.

### Availability of data and materials

After the clinical trial is finished, the original data will be uploaded to the ResMan Primitive Data Sharing Platform (IPD Sharing Platform) of the China Clinical Trials Registry at http://www.medresman.org:22280/login.aspx.

### Authors’ contributions

JF and ZHL initiated the study design. YNG and JH prepared the consent form.

YNG, JF and XYH drafted and finalized the study protocol.

All authors reviewed the study protocol and approved the final manuscript.

## Acknowledgements

The authors gratefully acknowledge the financial supports by Beijing Municipal Science&Technology Commission (Z171100001017066), and High Level Health Technical Talent Training Program of Beijing Municipal Health Bureau.

## Authors’ information

Beijing Tongren Eye Center, Beijing Tongren Hospital, Capital Medical University, Beijing Key Laboratory of Ophthalmology&Visual Science, Beijing, China

## References

1. Multi-ethnic Pediatric Eye Disease Study G. Prevalence of amblyopia and strabismus in African American and Hispanic children ages 6 to 72 months the multi-ethnic pediatric eye disease study. Ophthalmology 2008;115(7):1229–36 e1. doi: 10.1016/j.ophtha.2007.08.001 [published Online First: 2007/10/24]

2. McKean-Cowdin R, Cotter SA, Tarczy-Hornoch K, et al. Prevalence of amblyopia or strabismus in asian and non-Hispanic white preschool children: multi-ethnic pediatric eye disease study. Ophthalmology 2013;120(10):2117–24. doi: 10.1016/j.ophtha.2013.03.001 [published Online First: 2013/05/24]

3. Friedman DS, Repka MX, Katz J, et al. Prevalence of amblyopia and strabismus in white and African American children aged 6 through 71 months the Baltimore Pediatric Eye Disease Study. Ophthalmology 2009;116(11):2128–34 e1-2. doi: 10.1016/j.ophtha.2009.04.034 [published Online First: 2009/09/19]

4. Chia A, Dirani M, Chan YH, et al. Prevalence of amblyopia and strabismus in young singaporean chinese children. Invest Ophthalmol Vis Sci 2010;51(7):3411–7. doi: 10.1167/iovs.09-4461 [published Online First: 2010/03/09]

5. Hashemi H, Pakzad R, Heydarian S, et al. Global and regional prevalence of strabismus: a comprehensive systematic review and meta-analysis. Strabismus 2019;27(2):54–65. doi: 10.1080/09273972.2019.1604773 [published Online First: 2019/04/24]

6. CW P, H Z, JJ Y, et al. Epidemiology of Intermittent Exotropia in Preschool Children in China. Optometry and vision science : official publication of the American Academy of Optometry 2016;93(1):57–62. doi: 10.1097/opx.0000000000000754

7. Fu J, Li SM, Liu LR, et al. Prevalence of amblyopia and strabismus in a population of 7th-grade junior high school students in Central China: the Anyang Childhood Eye Study (ACES). Ophthalmic Epidemiol 2014;21(3):197–203. doi: 10.3109/09286586.2014.904371 [published Online First: 2014/04/20]

8. Yang CQ, Shen Y, Gu YS, et al. Clinical investigation of surgery for intermittent exotropia. J Zhejiang Univ Sci B 2008;9(6):470–3. doi: 10.1631/jzus.B0720007 [published Online First: 2008/06/11]

9. Bruce A, Santorelli G. Prevalence and Risk Factors of Strabismus in a UK Multi-ethnic Birth Cohort. Strabismus 2016;24(4):153–60. doi: 10.1080/09273972.2016.1242639 [published Online First: 2016/12/09]

10. Nusz KJ, Mohney BG, Diehl NN. The Course of Intermittent Exotropia in a Population-Based Cohort. Ophthalmology 2006;113(7):1154–58. doi: 10.1016/j.ophtha.2006.01.033

11. Govindan M, Mohney BG, Diehl NN, et al. Incidence and types of childhood exotropia: a population-based study. Ophthalmology 2005;112(1):104–8. doi: 10.1016/j.ophtha.2004.07.033 [published Online First: 2005/01/05]

12. Wallace DK, Christiansen SP, Sprunger DT, et al. Esotropia and Exotropia Preferred Practice Pattern(R). Ophthalmology 2018;125(1):P143–P83. doi: 10.1016/j.ophtha.2017.10.007 [published Online First: 2017/11/08]

13. Yao J, Wang X, Ren H, et al. Ultrastructure of medial rectus muscles in patients with intermittent exotropia. Eye (Lond) 2016;30(1):146–51. doi: 10.1038/eye.2015.213 [published Online First: 2015/10/31]

14. Bui Quoc E, Milleret C. Origins of strabismus and loss of binocular vision. Front Integr Neurosci 2014;8:71. doi: 10.3389/fnint.2014.00071 [published Online First: 2014/10/14]

15. Ahn SJ, Yang HK, Hwang JM. Binocular visual acuity in intermittent exotropia: role of accommodative convergence. Am J Ophthalmol 2012;154(6):981–86 e3. doi: 10.1016/j.ajo.2012.05.026 [published Online First: 2012/09/11]

16. Hatt SR, Leske DA, Mohney BG, et al. Fusional convergence in childhood intermittent exotropia. Am J Ophthalmol 2011;152(2):314–9. doi: 10.1016/j.ajo.2011.01.042 [published Online First: 2011/05/31]

17. Brodsky MC, Jung J. Intermittent Exotropia and Accommodative Esotropia: Distinct Disorders or Two Ends of a Spectrum? Ophthalmology 2015;122(8):1543–6. doi: 10.1016/j.ophtha.2015.03.004 [published Online First: 2015/07/27]

18. Hatt SR, Leske DA, Adams WE, et al. Quality of life in intermittent exotropia: child and parent concerns. Arch Ophthalmol 2008;126(11):1525–9. doi: 10.1001/archopht.126.11.1525 [published Online First: 2008/11/13]

19. Wang Y, Xu M, Yu H, et al. Health-related quality of life correlated with the clinical severity of intermittent exotropia in children. Eye (Lond) 2019;34(2):400–407 doi: 10.1038/s41433-019-0557-1 [published Online First: 2019/08/14]

20. RS H, EL S, ML C. Motor and sensory fusion in monkeys: psychophysical measurements. Eye (London, England) 1996:209–16. doi: 10.1038/eye.1996.48

21. Yang X, Zhang J, Lang L, et al. Assessment of cortical dysfunction in infantile esotropia using fMRI. Eur J Ophthalmol 2014;24(3):409–16. doi: 10.5301/ejo.5000368 [published Online First: 2013/10/31]

22. Feng X, Zhang X, Jia Y. Improvement in fusion and stereopsis following surgery for intermittent exotropia. J Pediatr Ophthalmol Strabismus 2015;52(1):52–7. doi: 10.3928/01913913-20141230-08 [published Online First: 2015/02/03]

23. G T, X H, Y Z, et al. A functional MRI study of altered spontaneous brain activity pattern in patients with congenital comitant strabismus using amplitude of low-frequency fluctuation. Neuropsychiatric disease and treatment 2016;12:1243–50. doi: 10.2147/ndt.S104756

24. Yan X, Lin X, Wang Q, et al. Dorsal visual pathway changes in patients with comitant extropia. PLoS One 2010;5(6):e10931. doi: 10.1371/journal.pone.0010931 [published Online First: 2010/06/10]

25. Li Q, Bai J, Zhang J, et al. Assessment of Cortical Dysfunction in Patients with Intermittent Exotropia: An fMRI Study. PLoS One 2016;11(8):e0160806. doi: 10.1371/journal.pone.0160806 [published Online First: 2016/08/09]

26. A M, JC H, GT L. Functional magnetic resonance imaging and its clinical utility in patients with visual disturbances. Survey of ophthalmology 2002;47(6):562–79. doi: 10.1016/s0039-6257(02)00356-9

27. Chen X, Zirnsak M, Vega GM, et al. Parietal Cortex Regulates Visual Salience and Salience-Driven Behavior. Neuron 2020 doi: 10.1016/j.neuron.2020.01.016

28. Jamadar SD, Li S, Sforazzini F, et al. Simultaneous task-based BOLD-fMRI and [18-F] FDG functional PET for measurement of neuronal metabolism in the human visual cortex. Neuroimage 2019;189:258–66. doi: 10.1016/j.neuroimage.2019.01.003 [published Online First: 2019/01/08]

29. Morrison D, McSwain W, Donahue S. Comparison of sensory outcomes in patients with monofixation versus bifoveal fusion after surgery for intermittent exotropia. J AAPOS 2010;14(1):47–51. doi: 10.1016/j.jaapos.2009.11.015 [published Online First: 2010/03/17]

30. G T, ZR D, Y Z, et al. Altered brain network centrality in patients with adult comitant exotropia strabismus: A resting-state fMRI study. The Journal of international medical research 2018;46(1):392–402. doi: 10.1177/0300060517715340

31. D L, S L, X Z. Analysis of alterations in white matter integrity of adult patients with comitant exotropia. The Journal of international medical research 2018;46(5):1963–72. doi: 10.1177/0300060518763704

32. X Y, Y W, L X, et al. Altered Functional Connectivity of the Primary Visual Cortex in Adult Comitant Strabismus: A Resting-State Functional MRI Study. Current eye research 2019;44(3):316–23. doi: 10.1080/02713683.2018.1540642

33. J O, L Y, X H, et al. The atrophy of white and gray matter volume in patients with comitant strabismus: Evidence from a voxel-based morphometry study. Molecular medicine reports 2017;16(3):3276–82. doi: 10.3892/mmr.2017.7006

34. Zhu PW, Huang X, Ye L, et al. Altered intrinsic functional connectivity of the primary visual cortex in youth patients with comitant exotropia: a resting state fMRI study. Int J Ophthalmol 2018;11(4):668–73. doi: 10.18240/ijo.2018.04.22 [published Online First: 2018/04/21]

35. Shi H, Wang Y, Liu X, et al. Cortical Alterations by the Abnormal Visual Experience beyond the Critical Period: A Resting-state fMRI Study on Constant Exotropia. Curr Eye Res 2019:1–7. doi: 10.1080/02713683.2019.1639767 [published Online First: 2019/07/10]

36. Margines JB, Huang C, Young A, et al. Refractive Errors and Amblyopia Among Children Screened by the UCLA Preschool Vision Program in Los Angeles County. Am J Ophthalmol 2020;210:78–85. doi: 10.1016/j.ajo.2019.10.013 [published Online First: 2019/10/28]

37. Mills MD, Coats DK, Donahue SP, et al. Strabismus surgery for adults: a report by the American Academy of Ophthalmology. Ophthalmology 2004;111(6):1255–62. doi: 10.1016/j.ophtha.2004.03.013 [published Online First: 2004/06/05]

38. Buck D, Clarke MP, Haggerty H, et al. Grading the severity of intermittent distance exotropia: the revised Newcastle Control Score. Br J Ophthalmol 2008;92(4):577. doi: 10.1136/bjo.2007.120287 [published Online First: 2008/03/29]

39. C W. Squint, Its Causes, Pathology, and Treatment. Edinburgh Medical Journal 1903;14(4):350–50.

40. Cox MA, Dougherty K, Westerberg JA, et al. Temporal dynamics of binocular integration in primary visual cortex. J Vis 2019;19(12):13. doi: 10.1167/19.12.13 [published Online First: 2019/10/18]

41. Grossberg S, Srinivasan K, Yazdanbakhsh A. Binocular fusion and invariant category learning due to predictive remapping during scanning of a depthful scene with eye movements. Front Psychol 2014;5:1457. doi: 10.3389/fpsyg.2014.01457 [published Online First: 2015/02/03]

42. AR OC, EE B, S A, et al. Relationship between binocular vision, visual acuity, and fine motor skills. Optometry and vision science : official publication of the American Academy of Optometry 2010;87(12):942–7. doi: 10.1097/OPX.0b013e3181fd132e

43. P F, RS H. The relative sensitivities of sensory and motor fusion to small binocular disparities. Vision research 2001;41(15):1969–79. doi: 10.1016/s0042-6989(01)00081-5

44. Economides JR, Adams DL, Horton JC. Capturing the Moment of Fusion Loss in Intermittent Exotropia. Ophthalmology 2017;124(4):496–504. doi: 10.1016/j.ophtha.2016.11.039 [published Online First: 2017/01/14]

45. Dickmann A, Aliberti S, Rebecchi MT, et al. Improved sensory status and quality-of-life measures in adult patients after strabismus surgery. J AAPOS 2013;17(1):25–8. doi: 10.1016/j.jaapos.2012.09.017 [published Online First: 2013/01/29]

46. MB M, C B, BA H. Binocularity following surgical correction of strabismus in adults. Journal of AAPOS : the official publication of the American Association for Pediatric Ophthalmology and Strabismus 2004;8(5):435–8. doi: 10.1016/j.jaapos.2004.07.003

47. Morris RJ, Scott WE, Dickey CF. Fusion after Surgical Alignment of Longstanding Strabismus in Adults. Ophthalmology 1993;100(1):135–38. doi: 10.1016/s0161-6420(93)31703-3

48. F K, Y E, NS Y. Does restoration of binocular vision make any difference in the quality of life in adult strabismus. The British journal of ophthalmology 2013;97(11):1425–30. doi: 10.1136/bjophthalmol-2013-303704

49. Chan ST, Tang KW, Lam KC, et al. Neuroanatomy of adult strabismus: a voxel-based morphometric analysis of magnetic resonance structural scans. Neuroimage 2004;22(2):986–94. doi: 10.1016/j.neuroimage.2004.02.021 [published Online First: 2004/06/15]

50. Shao Y, Li QH, Li B, et al. Altered brain activity in patients with strabismus and amblyopia detected by analysis of regional homogeneity: A restingstate functional magnetic resonance imaging study. Mol Med Rep 2019;19(6):4832–40. doi: 10.3892/mmr.2019.10147 [published Online First: 2019/05/07]

51. D C, D P. The extraordinarily rapid disappearance of entoptic images. Proceedings of the National Academy of Sciences of the United States of America 1996;93(15):8001–4. doi: 10.1073/pnas.93.15.8001

